# CTCA-Based Pericoronary Fat and Anatomy–Flow Signatures Identify Future Culprit Lesions in Moderate Stenoses

**DOI:** 10.64898/2026.05.12.26352858

**Authors:** Mingzi Zhang, Chi Shen, Lucy McGrath-Cadell, Ramtin Gharleghi, Hassan Assareh, James Otton, Daniel A. Moses, Jolanda J. Wentzel, Robert M. Graham, Craig S. McLachlan, Susann Beier

## Abstract

**Background:** A substantial proportion of coronary events originate from angiographically moderate lesions, indicating that stenosis severity alone does not reflect lesion biomechanical risk.

**Objectives:** To test whether adding lesion-adjacent pericoronary adipose tissue (PCAT) and CTCA-derived anatomy–flow descriptors to quantitative plaque assessment improves identification of future culprit lesions, with a prespecified focus on moderate stenosis.

**Methods:** We performed a within-patient, lesion-level case–control analysis in the GeoCAD cohort, including patients undergoing coronary revascularisation during follow-up. Culprit lesions were identified from longitudinal CTCA. Stenosis severity, quantitative plaque composition, and PCAT volume were quantified (MEDIS), and vessel centreline geometry and lesion haemodynamics derived using computational modelling. Incremental prognostic value was assessed using Cox models with drop-one and stepwise workflow analyses, including a prespecified subgroup analysis of moderate stenosis lesions (25–49% diameter stenosis).

**Results:** Among 46 patients (212 lesions; 55 culprit), percent area stenosis (%AS) dominated culprit lesion discrimination (HR: 2.01; 95% CI: 1.54–2.62; p<0.001). In 82 moderate-stenosis lesions (30 culprit), %AS provided minimal discrimination (ΔC-index: 0.01; p=0.895). Culprit lesions were characterised by greater PCAT volume (HR: 1.75; 95% CI: 1.29–2.37; p<0.001), higher helical flow intensity (HR: 1.35; 95% CI: 1.16–1.57; p<0.001), and lower torsion (HR: 0.50; 95% CI: 0.29–0.84; p=0.009). Adding anatomy– flow descriptors improved risk stratification for moderate lesions beyond CTCA stenosis and plaque/PCAT features (p=0.007).

**Conclusions:** In moderate stenosis, lesion-adjacent PCAT and anatomy–flow descriptors provided incremental prognostic information beyond luminal narrowing and plaque composition, supporting integrated CTCA phenotyping to identify high-risk nonobstructive coronary lesions.

**CONDENSED ABSTRACT:** A substantial proportion of coronary events arise from nonobstructive, limiting stenosis-based risk assessment on CTCA. In a lesion-level analysis of the GeoCAD cohort, we tested whether adding lesion-adjacent pericoronary adipose tissue (PCAT) and CTCA-derived anatomy–flow descriptors improves identification of culprit lesions. Among 212 lesions, stenosis severity dominated discrimination overall but showed minimal value in 82 moderately stenotic lesions (25–49% diameter stenosis). In this subgroup, PCAT volume, helical flow intensity, and centreline torsion provided stronger prognostic value, and their integration improved risk stratification beyond CTCA stenosis and plaque features, supporting integrated CT phenotyping of high-risk nonobstructive lesions.

## Introduction

Coronary computed tomography angiogram (CTCA) provides robust assessment of luminal stenosis, plaque burden, and recognised adverse plaque features, yet predicting which individual coronary lesions will subsequently require clinically indicated revascularisation remains imperfect. Importantly, longitudinal natural-history studies have demonstrated that a substantial proportion of future coronary events and revascularisations arise from lesions that are non-obstructive or only moderately stenotic, highlighting the limitations of a purely stenosis-centric paradigm.^1,2^.

Importantly, much of the imaging and interventional literature has focused on angiographically intermediate lesions, with the primary aim of identifying haemodynamically significant disease suitable for revascularisation. Physiology-guided strategies, such as fractional flow reserve (FFR), have proven effective for this purpose and underpin contemporary clinical practice ^3^. In parallel, CTCA-based studies have associated qualitative vulnerable plaque features, including the napkin-ring sign, low-attenuation plaque, positive remodelling, and spotty calcification, with subsequent events ^4^. Additional work has linked surface-derived endothelial shear stress (ESS) metrics to plaque progression and myocardial infarction ^5,6^, while pericoronary adipose tissue (PCAT) attenuation has emerged as a surrogate for local vascular inflammation with incremental prognostic value ^7,8^.

Despite these advances in examining stress and haemodynamic factor superimposed on coronary lesions assessments, important gaps remain. Most prior studies emphasise vessel-or patient-level ischaemic risk rather than lesion-level prediction in earlier-stage, non–flow-limiting disease, particularly within the moderate-stenosis range where lesions are prevalent and clinically consequential. Moreover, many CTCA studies rely on visual or semi-quantitative plaque descriptors and surface-based haemodynamic indices, while volumetric flow patterns and local centreline geometry—key determinants of three-dimensional flow organisation and near-wall transport ^9^—are less frequently integrated. Recent expert statements have underscored the importance of quantitative plaque assessment for advancing CT-based risk stratification ^10^, however also highlighting substantial between-study heterogeneity and the need for standardised, lesion-level approaches that identify future risk ^11^.

Accordingly, in this within-patient, lesion-level case–control analysis from the GeoCAD cohort ^12^, we evaluated whether integrating CTCA-derived anatomical severity, quantitative plaque composition, lesion-adjacent PCAT metrics, centreline geometry, and computationally derived flow patterns improves lesion-level prediction of subsequent clinically indicated revascularisation. We further prespecified a subgroup analysis in lesions with moderate stenosis (25–49% diameter stenosis), where clinical decision-making is frequently uncertain and where improved lesion-level risk stratification may be most valuable for preventive management.

## 2. Methods

### 2.1 Study population

This study was a lesion-level case-control analysis derived from the GeoCAD cohort^12^, a retrospective study of longitudinal CT coronary angiograms (CTCA) and clinical follow-up. For the present analysis, we included only patients who underwent clinical indicated coronary revascularisation during follow-up, enabling lesion-level attribution of major adverse cardiac events (MACE). **Figure 1** presents the study flow diagram for patient inclusion and exclusion. Culprit lesions were identified by locating stents on post-revascularisation CTCA imaging; when no post-procedure CTCA was available, two experienced cardiologists (LMC and JO) independently reviewed the longitudinal imaging series, using the ROMICAT (Rule Out Myocardial Infarction by Computer Assisted Tomography) definitions ^13^, to determine culprit lesions by consensus. All analyses were performed using images acquired prior to revascularisation. Because revascularisation is influenced by clinical and procedural factors, this endpoint reflects clinically driven treatment rather than spontaneous plaque rupture; however, the within-patient design enables robust lesion-level comparisons of incremental imaging value.

**Figure 1.**
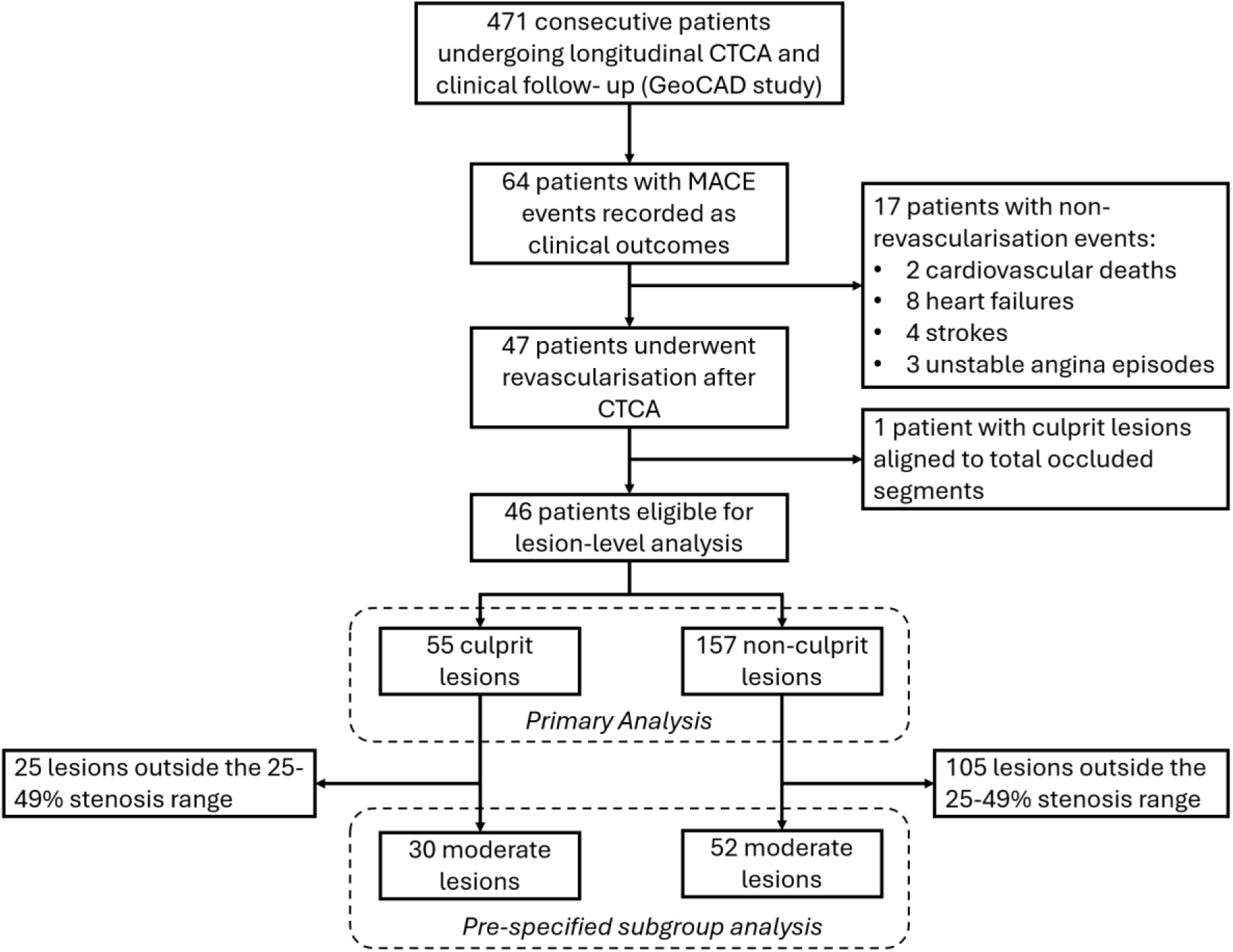
Patient inclusion diagram.

This study was approved by St Vincent’s Hospital Human Research Ethics Committee (2020/ETH02127) and the NSW Population and Health Service Research Ethics Committee (2021/ETH00990) and conducted in accordance with Declaration of Helsinki.

### 2.2 CTCA imaging and coronary anatomy reconstruction

The CTCA images were acquired using the multi-detector GE Revolution CT Scanners at Spectrum Medical Imaging, Sydney, using a prospective ECG-gated protocol. The scanning involved achieving a patient heart rate of around 60 beats per minute by administration of beta blockers. The patients then received sublingual nitrates and an intravenous injection of a bolus of iodine-based contrast agent before acquisition of the end-diastolic images.

We used a combined EfficientNet and LinkNet neural network ^14^ to prepare the coronary artery segmentations for manual verification for their good performance in capturing vessel boundaries. Automated coronary segmentations were then verified by experienced cardiologists (LMC and JO). The Vascular Modelling ToolKits (VMTK v1.4) was used to generate the coronary centrelines, where inlets and outlets were labelled and average maximal inscribe sphere diameters, curvatures, and torsions extracted that corresponded to discretised centreline points. Detailed definitions of the anatomic and blood flow metrics were in the **Supplemental Methods**.

### 2.3 Lesion characteristics, plaque components, and pericoronary adipose tissue

Following the radiological reports, two readers (MZ, CS) independently annotated the lesion throat, proximal and distal ends of a lesion using the MEDIS Medical Imaging System QAngio CT (v2025.1.14.6). All annotations were subsequently adjudicated by a third senior expert (LMC), and discrepancies were resolved by consensus. From these annotations, lesions-level quantitative metrics, including lesion length, plaque volume, plaque burden, minimal lumen area, remodelling index, plaque eccentricity, and percent diameter and area stenosis (%DS and %AS), were derived.

In addition to total plaque volume, the plaque composition was quantified semi-automatically within MEDIS, including the proportional volumes of calcified plaque (≥350 HU), fibrous plaque (131-350 HU), fibrofatty plaque (31-130 HU), and necrotic core (−30 to 30 HU)^15^. Pericoronary adipose tissue (PCAT) was quantified using the same software platform. PCAT intensity and volume were measured within 3 mm distance from the outer coronary vessel wall, following established thresholds for adipose tissue in the range from −190 to −30 HU ^8^. These metrics were extracted to capture local pericoronary inflammatory signatures relevant to coronary lesion activity.

### 2.4 Coronary blood flow patterns characterisation

Detailed computational procedures are provided in the **Supplemental Methods**. In brief, coronary flow simulations were performed using an automated pipeline built around the Python-based CadToolKit library, which manages coronary model preparation, boundary-condition assignment, and execution of the finite-volume CFD solver (CFX, ANSYS, v2023r1). Coronary geometries were pre-processed using standard vascular modelling utilities (VMTK, TriMesh, etc), and distal branches were truncated at a lumen diameter of 1.5 mm. Blood was modelled as an incompressible, non-Newtonian fluid using the Carreau-Yasuda viscosity model. Inflow conditions were adapted to each patient by scaling published coronary blood flow profiles according to left main and proximal right coronary artery diameters^16^, while outflow distribution was assigned using a morphometry-based scaling law, applied recursively at each bifurcation^17^.

Resting coronary haemodynamics were simulated over four cardiac cycles, and results from the final cycle were used for analysis to minimise transient start-up effects. Six haemodynamic descriptors were extracted on a per-lesion basis. Time-averaged endothelial shear stress (TA-ESS) was computed over the cardiac cycle, from which two burden measures were derived: LowESS, the %area exposed to TA-ESS<0.5 Pa, associated with plaque progression^6^, and HighESS, the %area exposed to TA-ESS>4.71 Pa, associated with plaque rupture^18^. Additional surface-based descriptors included TA-ESS gradient (TA-ESSG), quantifying the spatial shear heterogeneity relevant to plaque disruption^19^, and the topological shear variation index (TSVI), reflecting dynamic shear-induced endothelial expansion and contraction predictive of myocardial infarction^20^. The volume-based descriptor, average helical flow intensity (*h*_2_), was calculated to characterise the 3D complexity of flow patterns through stenotic segments^21^.

All metrics were sampled at the predefined locations along each lesion: (i) proximal end, (ii) throat, and (iii) distal end, each averaged across a 3-mm segment centred on the location, and additionally across longitudinal spans (proximal end to throat, proximal end to distal end, and throat to distal end) to characterise haemodynamic variation along the lesion. To capture not only focal values but also the overall spatial pattern of haemodynamics, principal component analysis (PCA) was performed across all location-specific measurements for each metric. This approach enabled the extraction of dominant modes of variation along the lesion length, reflecting how haemodynamic stress evolves from proximal to distal vessel segments. The first principal component (PC1), which explained the majority of spatial variance, was used as a surrogate descriptor of the lesion-level haemodynamic pattern for subsequent statistical analysis.

### 2.5 Statistical analysis

Lesion-level analyses were structured around two complementary objectives: (1) to evaluate the incremental prognostic contribution of distinct pathophysiological descriptors associated with coronary disease, and (2) to assess how risk prediction improves across a stepwise CTCA-based diagnostic workflow—from routine anatomical assessment on CTCA, to advanced quantitative plaque analysis using MEDIS, and finally to CFD-derived haemodynamic evaluation.

For the mechanistic analysis, candidate parameters were grouped *a priori* into six pathophysiological families: (1) lesion severity, (2) plaque composition, (3) PCAT, (4) vessel shear stress metrics, (5) volumetric flow patterns, and (6) vessel geometry. To reduce model complexity and mitigate overfitting in this modest-sized cohort, one representative metric per family was selected for multivariable Cox modelling based on effect size, statistical significance, and biological interpretability. All continuous variables were standardised prior to modelling, and hazard ratios (HRs) are reported per 1–SD increase. The independent contribution of each metric was assessed using a drop-one approach, quantifying the change in Harrell’s C-index (ΔC-index) with 95% confidence intervals (CI) derived from bootstrap resampling. Model discrimination (C-index and ΔC-index) were estimated using bootstrap resampling to reduce optimism.

To reflect a clinically realistic CT diagnostic pathway, three nested models were constructed comprising: (1) lesion severity parameters obtainable directly from standard CTCA interpretation, (2) additional plaque composition and PCAT features requiring semi-automated post-processing (e.g., using MEDIS), and (3) lesion assessment incorporating centreline-based geometric descriptors and CFD-derived haemodynamic metrics. Predicted risks from these nested models were used to construct cumulative risk curves, illustrating how lesions risk stratification improves with the sequential addition of information from advanced analyses using MEDIS and CFD. Given the retrospective design and sample size, these models should be viewed as risk-enrichment tools; formal calibration and clinically deployable thresholds require larger, externally validated cohorts.

Because multiple lesions were analysed per patient, Cox models were fitted with patient-level cluster-robust standard errors. Continuous variables were compared using two-sided t-tests or Mann–Whitney U tests, and categorical variables using χ² or Fisher exact tests, as appropriate. Multiple testing was controlled using the Benjamini–Hochberg false discovery rate. A prespecified subgroup analysis was performed in lesions with moderate stenosis (%DS 25–49), given their numerical predominance and clinical relevance. All analyses were conducted in Python, with p<0.05 considered statistically significant after correction unless otherwise specified.

## 3. Results

### 3.1 Baseline characteristics of patients and lesions

A total of 46 patients (83% male, mean age 61.5±8.6 years) undergoing coronary revascularisation were included in this lesion-level case-control study, contributing 212 lesions (55 culprit and 157 non-culprit; **Figure 1**). The median Calcium score was 211 (IQR: 11–508) and modified Gensini score was 65.5±43.8. Most culprit lesions were of moderate stenosis (n=30/55, 54.5%), whereas non-culprit lesions were associated with mild stenosis (n=87/157, 55.4%). The moderate-stenosis subgroup comprised 82 lesions (30 culprit, 52 non-culprit). Median follow-up from imaging to revascularisation was 4.3 months overall and 4.1 months in the moderate subgroup. Additional characteristics are presented in **Tables 1** and **2**.

**Table 1.** Patient baseline characteristics.

| Characteristics | Patients (N = 46) |
| --- | --- |
| **Demographics** |  |
| Age, years | 61.5 ± 8.6 |
| Male | 38 (82.6%) |
| Height, cm | 170.2 ± 9.7 |
| Weight, kg | 82.3 ± 16.0 |
| BMI | 27.3 (25.1–31.8) |
| Heart Rate, bpm | 58.4 ± 8.5 |
| Systolic Blood Pressure, mmHg | 136.4 ± 22.2 |
| Diastolic Blood Pressure, mmHg | 79.5 ± 9.4 |
| Time to event, months |  |
| Overall | 4.32 (0.84–27.81) |
| Moderate-Stenosis | 4.14 (0.57–11.94) |
| **Risk Factors** |  |
| Obese | 12 (30.0%) |
| Hypertension | 26 (59.1%) |
| Dyslipidaemia | 29 (64.4%) |
| Diabetes | 12 (26.1%) |
| Smoker | 10 (26.3%) |
| Kidney Disease | 1 (2.2%) |
| Asthma | 1 (2.2%) |
| Family History | 8 (18.6%) |
| Calcium Score | 211.0 (10.8–507.8) |
| <100 | 17 (37.0%) |
| 100–399 | 12 (26.1%) |
| ≥400 | 17 (37.0%) |
| Modified Gensini Score | 65.5 ± 43.8 |
| <25 | 9 (19.6%) |
| 25–54 | 10 (21.7%) |
| ≥55 | 27 (58.7%) |
| **Medications** |  |
| OHAs | 10 (22.2%) |
| Insulin | 1 (2.2%) |
| Statin | 17 (37.8%) |
| ACE-I/ARB | 20 (44.4%) |
| Aspirin | 9 (20.0%) |
| P2Y12 inhibitors | 3 (6.7%) |
| CCB | 7 (15.6%) |
| Beta blocker | 10 (22.2%) |
| NOAC | 1 (2.2%) |
| Warfarin | 0 (0.0%) |

Note: Values are reported as counts (%) for categorical variables, mean ± standard deviations for normally distributed, or mean (interquartile ranges) for non-normally distributed continuous variables. Abbreviations: OHAs, oral hypoglycaemic agents; ACE-I/ARB, angiotensin-converting enzyme inhibitors/angiotensin receptor blockers; CCB, calcium channel blockers; NOAC, non-vitamin K antagonist oral anticoagulants.

**Table 2.**
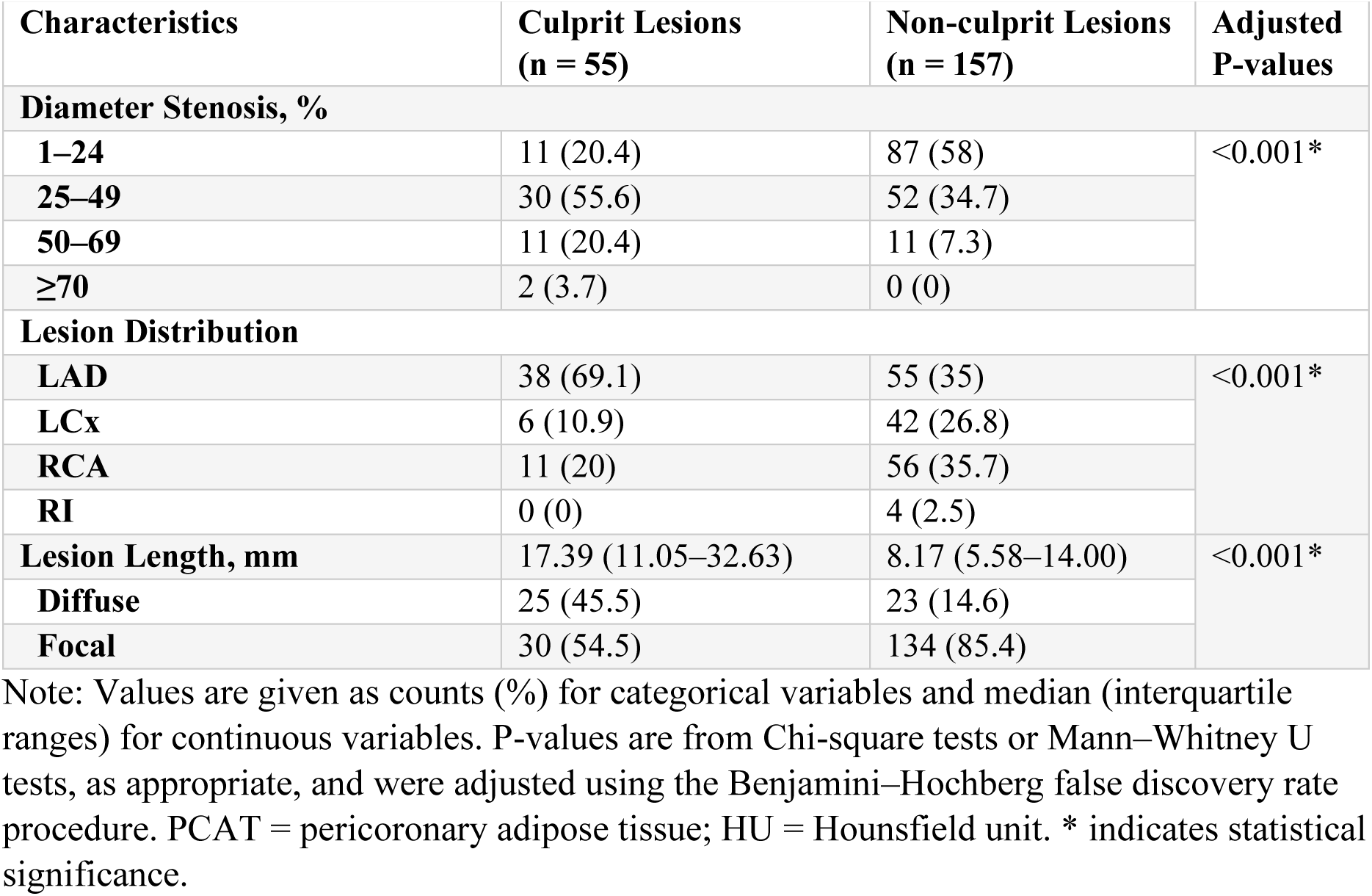
Lesion baseline characteristics.

| Characteristics | Culprit Lesions<br>(n = 55) | Non-culprit Lesions<br>(n = 157) | Adjusted<br>P-values |
| --- | --- | --- | --- |
| Diameter Stenosis, % |  |  |  |
| 1–24 | 11 (20.4) | 87 (58) | <0.001* |
| 25–49 | 30 (55.6) | 52 (34.7) |  |
| 50–69 | 11 (20.4) | 11 (7.3) |  |
| ≥70 | 2 (3.7) | 0 (0) |  |
| Lesion Distribution |  |  |  |
| LAD | 38 (69.1) | 55 (35) | <0.001* |
| LCx | 6 (10.9) | 42 (26.8) |  |
| RCA | 11 (20) | 56 (35.7) |  |
| RI | 0 (0) | 4 (2.5) |  |
| Lesion Length, mm | 17.39 (11.05–32.63) | 8.17 (5.58–14.00) | <0.001* |
| Diffuse | 25 (45.5) | 23 (14.6) |  |
| Focal | 30 (54.5) | 134 (85.4) |  |

Note: Values are given as counts (%) for categorical variables and median (interquartile ranges) for continuous variables. P-values are from Chi-square tests or Mann–Whitney U tests, as appropriate, and were adjusted using the Benjamini–Hochberg false discovery rate procedure. PCAT = pericoronary adipose tissue; HU = Hounsfield unit. \* indicates statistical significance.

### 3.2 Univariate analysis and feature selection for multivariate regression

Overall, culprit lesions had a larger plaque burden (0.43 vs 0.38; HR=1.38, p=0.02), plaque volume (66.4 vs 31.7 mm^3^; HR=1.49, p<0.001), and greater %AS (62 vs 37; HR=2.60, p<0.001). Positive remodelling was observed in 14 (25.5%) culprit lesions and 68 (43.3%) non-culprit lesions (p=0.037). With the highest HR, %AS was selected as the representative CTCA feature for multivariate modelling. Culprit lesions had significantly larger absolute volumes of the fibro-fatty, fibrous, calcium, and necrotic core components (all p<0.001). However, when considering the compositional percentage (%), only necrotic core (%NC) was statistically larger (8.1 vs 5.4, p=0.016) and therefore selected for the multivariate modelling. For pericoronary inflammation marker, PCAT volume (mm^3^) was selected, due to it being >2.5 fold larger in the culprit lesions (830 vs 317; HR=1.51, p<0.001). (**Table 3**) Lesion-specific pattern variations in the haemodynamic and anatomic characteristics were compared by their standardised non-dimensional first principal component (PC1). For ESS and its derivatives, both TSVI (1.16 vs –0.88; HR=1.50, p<0.001) and HighESS (0.08 vs – 1.27; HR=1.32, p=0.006) were larger in the culprit lesions. For the greater HR and its reported superiority over ESS in predicting culprit lesions ^20^, TSVI was selected as the representative surface-based haemodynamic predictor. Helical flow intensity (*h*_2_) was statistically higher in the culprit segments (–0.27 vs –0.34, p=0.004, **Supplementary Table S1**) and was the only volume-based haemodynamic metric to be included in the multivariate analysis. Vessels harbouring culprit lesions had a smaller standardised torsion (–0.85 vs –0.30; HR=0.64, p=0.024), but a greater curvature (0.10 vs –0.48; HR=1.15, p=0.064), with torsion selected for multivariate regression. (**Table 4**)

**Table 3.** MEDIS-derived CTCA lesion characteristics and standardised Cox regression in overall and moderate-stenosis lesions.

| Variables | All Lesions |  |  |  | Moderate-Stenosis Lesions |  |  |  |
| --- | --- | --- | --- | --- | --- | --- | --- | --- |
|  | Culprit<br>(n = 55) | Non-culprit<br>(n = 157) | HR (95% CI) | Wald test<br>for HR | Culprit<br>(n = 30) | Non-culprit<br>(n = 52) | HR (95% CI) | Wald test<br>for HR |
| **Plaque Burden** | 0.43 (0.36–0.50) | 0.38 (0.31–0.45) | 1.38 (1.05, 1.80) | 0.02* | 0.38 (0.33–0.48) | 0.42 (0.34–0.49) | 1.04 (0.74, 1.46) | 0.807 |
| **Minimal Lumen Area, mm<sup>2</sup>** | 3.12 (1.81–4.56) | 4.61 (2.79–7.02) | 0.49 (0.39, 0.62) | <0.001* | 3.22 (2.32–4.16) | 3.33 (2.05–4.66) | 0.73 (0.53, 0.99) | 0.043* |
| **Plaque Eccentricity** | 0.9 (0.8–1.0) | 1.0 (0.9–1.0) | 0.67 (0.54, 0.82) | <0.001* | 0.9 (0.9–1.0) | 1.0 (0.9–1.0) | 0.91 (0.70, 1.18) | 0.466 |
| **Area Stenosis** | 0.6 (0.5–0.7) | 0.3 (0.2–0.6) | 2.60 (2.03, 3.33) | <0.001* | 0.6 (0.5–0.7) | 0.6 (0.5–0.7) | 1.32 (0.90, 1.96) | 0.158 |
| **Positive Remodelling** | 14 (25.5%) | 68 (43.3%) | 0.55 (0.32, 0.97) | 0.037* | 4 (13.3%) | 12 (23.1%) | 0.80 (0.25, 2.58) | 0.707 |
| **Plaque Volume, mm<sup>3</sup>** | 66.4 (38.2–175.2) | 31.7 (17.7–68.4) | 1.49 (1.27, 1.74) | <0.001* | 71.5 (40.8–139.3) | 33.6 (20.6–92.2) | 1.41 (1.12, 1.78) | 0.003* |
| **PCAT Volume, mm<sup>3</sup>** | 829.9 (312.5–1713.2) | 317.2 (173.5–584.8) | 1.51 (1.27, 1.80) | <0.001* | 1021.4 (426.5–1528.7) | 348.3 (189.1–807.9) | 1.30 (1.00, 1.70) | 0.052 |
| **Mean PCAT Intensity, HU** | –80.7 (–84.8–<br>–75.7) | –77.4 (–82.7–<br>–70.8) | 0.75 (0.61, 0.93) | 0.009* | –80.9 (–85.1–<br>–77.7) | –77.3 (–84.5–<br>–73.1) | 0.99 (0.76, 1.30) | 0.937 |
| **Compositional Volumes, mm<sup>3</sup>** |  |  |  |  |  |  |  |  |
| **Fibrous** | 28.5 (14.6–77.3) | 15.6 (7.5–31.8) | 1.45 (1.25, 1.69) | <0.001* | 28.5 (20.4–73.5) | 18.4 (7.9–43.5) | 1.31 (0.98, 1.75) | 0.068 |
| **Fibro-fatty** | 12.0 (7.0–24.2) | 4.4 (2.3–9.0) | 1.81 (1.52, 2.16) | <0.001* | 17.6 (7.4–27.5) | 4.5 (2.4–11.2) | 1.80 (1.41, 2.30) | <0.001* |
| **Necrotic Core** | 9.9 (2.7–20.2) | 1.9 (0.6–4.8) | 1.55 (1.30, 1.86) | <0.001* | 9.9 (2.6–22.3) | 2.9 (0.6–8.6) | 1.45 (1.16, 1.82) | 0.001* |
| **Calcium** | 7.3 (0.3–57.2) | 5.8 (1.1–17.5) | 1.37 (1.18, 1.58) | <0.001* | 4.5 (0.1–23.0) | 7.1 (2.1–23.0) | 1.21 (0.87, 1.66) | 0.254 |
| **Compositional Percentages, %** |  |  |  |  |  |  |  |  |
| **Fibrous** | 41.9 (30.3–55.3) | 47.8 (37.2–61.9) | 0.81 (0.64, 1.01) | 0.065 | 43.5 (33.3–57.2) | 46.9 (37.1–58.7) | 0.88 (0.65, 1.17) | 0.374 |
| **Fibro-fatty** | 16.7 (9.3–24.9) | 14.7 (9.7–19.5) | 1.19 (0.83, 1.69) | 0.341 | 19.0 (12.9–27.2) | 14.8 (9.7–17.5) | 1.65 (1.19, 2.30) | 0.003* |
| **Necrotic Core** | 8.1 (3.8–17.3) | 5.4 (2.3–11.7) | 1.09 (0.85, 1.40) | 0.482 | 10.2 (4.7–16.4) | 6.7 (2.2–14.4) | 1.11 (0.85, 1.44) | 0.438 |
| **Calcium** | 17.0 (0.5–42.7) | 25.7 (8.7–37.3) | 1.05 (0.75, 1.47) | 0.763 | 8.3 (0.3–32.5) | 24.8 (12.8–37.1) | 0.83 (0.54, 1.28) | 0.398 |

Note: Values are median (Q1–Q3) or n (%), as appropriate. Hazard ratios (HRs) were derived from Cox proportional hazards models and are reported per 1–SD increase in each variable. P values are results of the Cox regression analyses, and \*indicates statistical significance. PCAT = pericoronary adipose tissue. Moderate stenosis was defined as a diameter stenosis of 25%–49%.

**Table 4.**
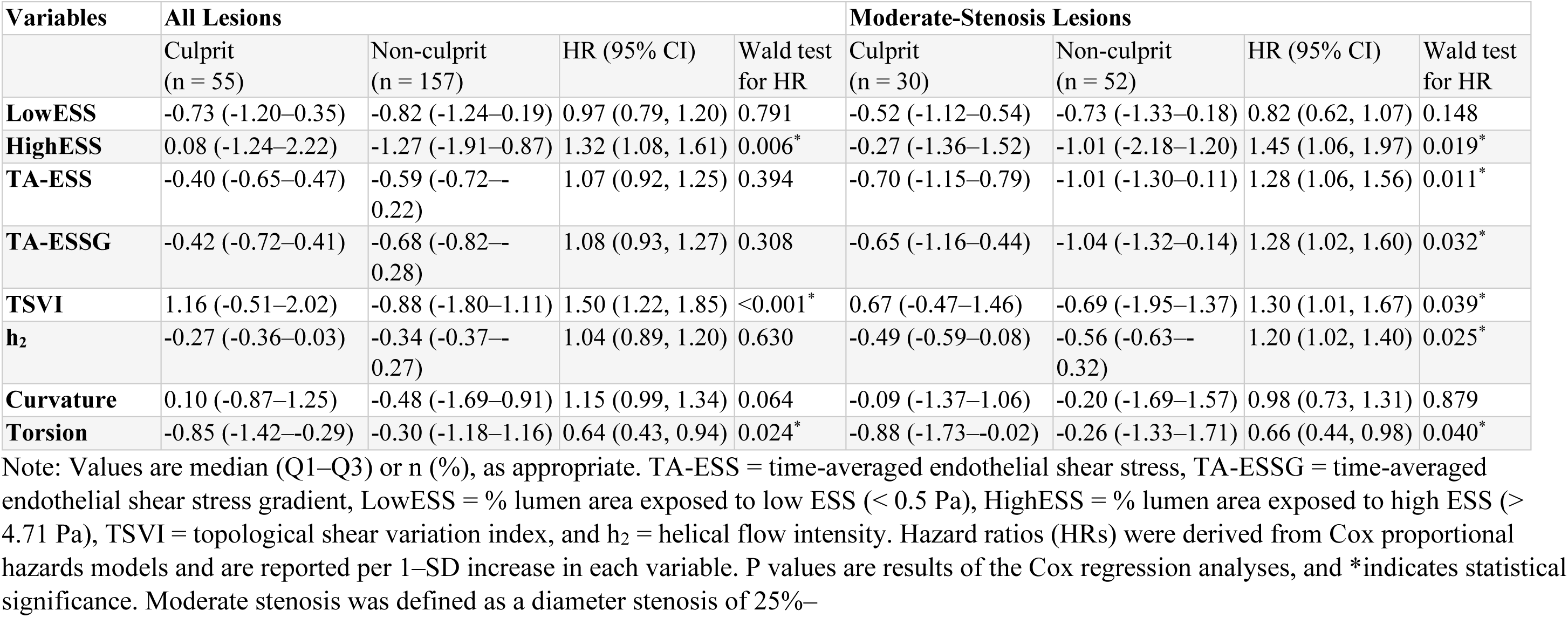
Lesion-specific anatomic and blood flow pattern variations, quantified as the first principal component (PC1), and standardised Cox regression in overall and moderate-stenosis lesions.

### 3.3 Multivariate culprit-lesion prediction model and the incremental value of each predictor

In the overall lesion group, multivariate prediction models, containing parameters of %AS, %NC, PCAT volume, TSVI, *h*_2_, and torsion, demonstrated stable discriminatory performance, with the highest Harrell’s C-index achieved by the full-feature model (C=0.80, 95% CI: 0.74–0.86). Drop-one tests showed that removal of %AS produced the largest decrement (ΔC=0.03, 95% CI: −0.00–0.09; p<0.001), indicating its contributing role to model performance in this cohort (**Figure 2, A**).

**Figure 2.**
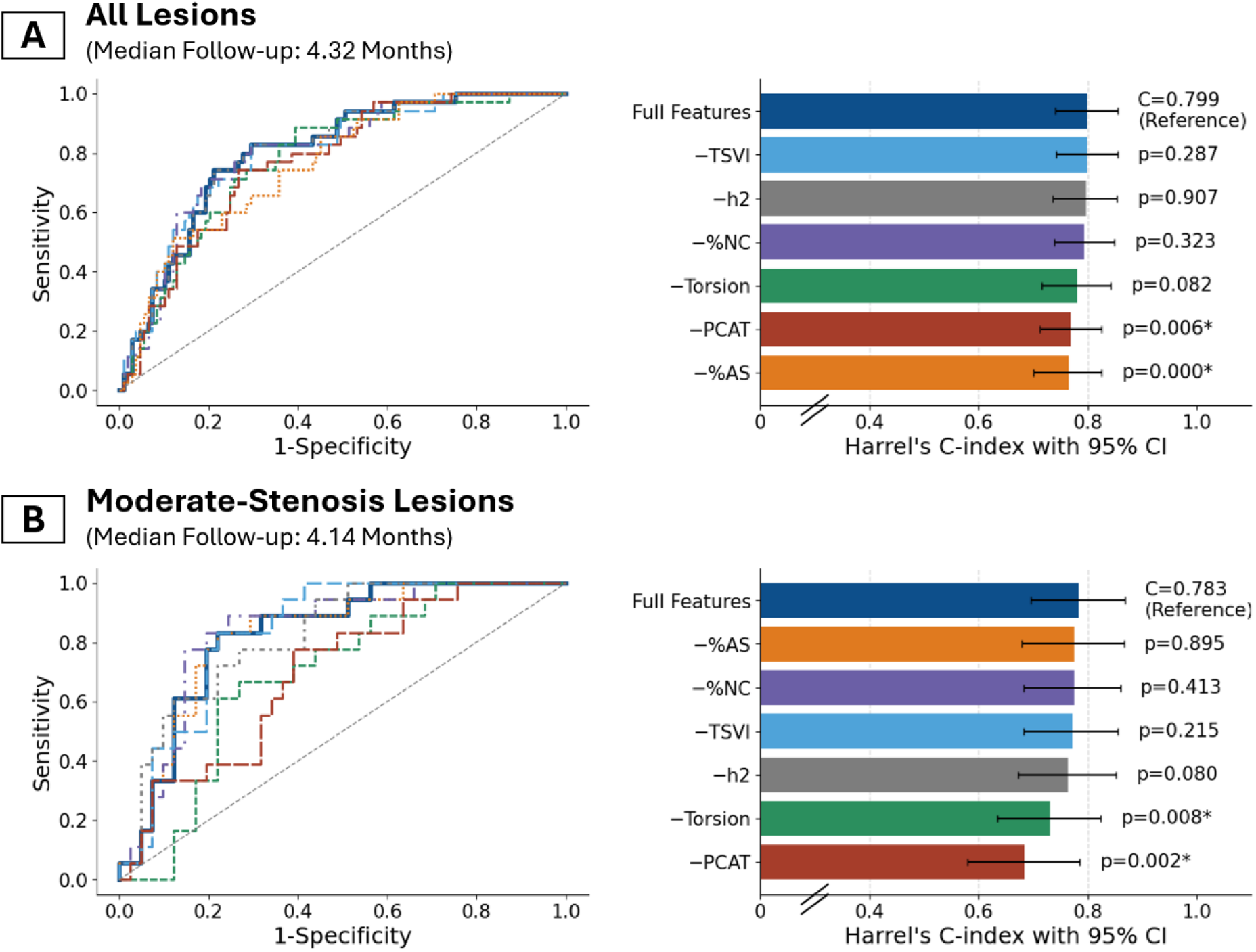
Drop-one analysis of lesion-level imaging features for culprit prediction. Receiver operating characteristic (ROC) curves (left) and Harrell’s C-index with 95% confidence intervals (right) show the incremental prognostic contribution of six lesion-specific imaging features in the overall lesion cohort (A) and in moderate-stenosis lesions (B). The full multivariable Cox model, including all features, served as the reference; each drop-one model sequentially excluded a single feature, and the resulting change in discrimination was quantified as ΔC-index. ROC curves are shown at the median follow-up. Abbreviations: TSVI=topological shear variation index; PCAT=pericoronary adipose tissue; h2=helical flow intensity.

However, discrimination in the moderate-stenosis subgroup was more sensitive to the removal of PCAT and lesion-specific haemodynamic and vessel anatomic features — dropping the PCAT volume led to a marked reduction in C-index (ΔC=0.10, 95% CI: 0.00– 0.21; p=0.002), followed by vessel torsion (ΔC=0.05, 95% CI: 0.00–0.13; p=0.008) and helical flow intensity (ΔC=0.02, 95% CI: −0.02–0.08; p=0.08), whereas removal of %AS had minimal impact on model discrimination (ΔC=0.01, 95% CI: −0.03–0.06; p=0.895; **Figure 2, B**).

Composite exposure and multivariate HR analyses further demonstrated cohort-specific patterns, with PCAT, haemodynamic, and vessel anatomic features driving progressive culprit–non-culprit separation only in moderate-stenosis lesions, whereas stenosis severity dominated discrimination in the overall cohort. Correspondingly, multivariable HR showed attenuation of associations for PCAT volume, *h*_2_, and vessel torsion in the overall cohort, while these associations were comparatively stronger in moderate-stenosis lesions after multivariable adjustment (**Figure 3**).

**Figure 3.**
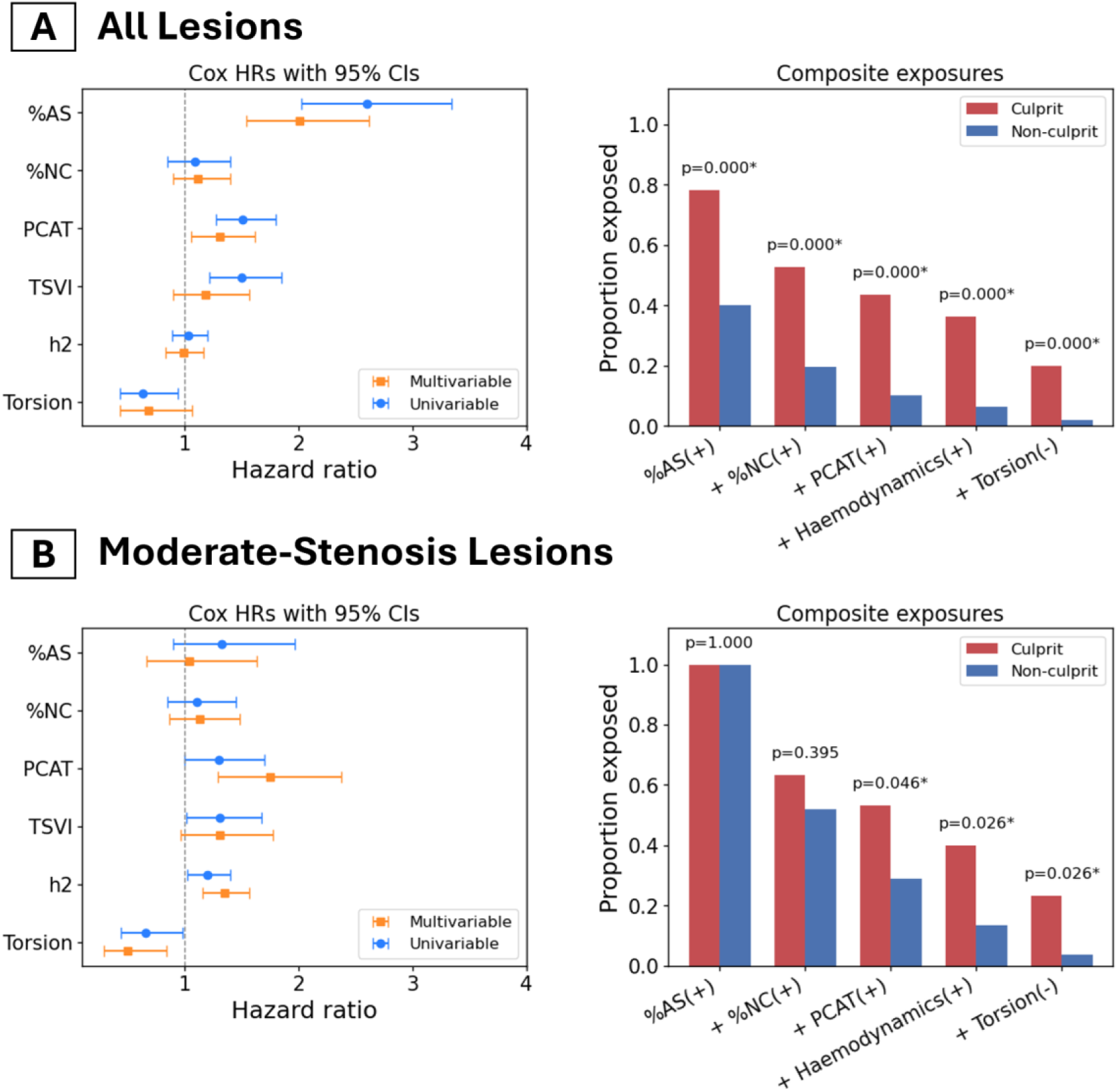
Composite exposure enrichment and hazard ratios for lesion-level culprit events in the overall cohort (A) and the moderate-stenosis subset (B). Left panels: proportions of lesions classified as progressively complex exposure patterns — (1) high area stenosis (%AS), (2) %AS + high necrotic core (%NC), (3) + high PCAT (pericoronary coronary adipose volume), (4) + high haemodynamics (topological shear variation index, TSVI, or h2), and (5) + low Torsion — shown separately for culprit (red) and non-culprit (blue) lesions. Binary thresholds were derived from cohort medians in the overall file. P-values above each bar pair are chi-square test after correction with the Benjamini–Hochberg FDR procedure. Right panels: univariable (blue circles) and multivariable (orange squares) Cox proportional hazards estimates for each predictor. Hazard ratios are reported per 1-SD (z-score) increase and error bars are 95% CIs. The dashed vertical line indicates HR=1.0.

### 3.4 Clinical relevance of stepwise CTCA-based diagnostic workflow

Predicted cumulative risk curves derived from nested CTCA-based models demonstrated distinct patterns of risk stratification across diagnostic strategies (i.e. CTCA, CTCA+MEDIS, or CTCA+MEDIS+CFD, **Figure 4**). In the overall lesions cohort, incorporation of MEDIS-derived PCAT and plaque component features resulted in a modest incremental separation of risk curves over follow-up (p=0.042). In contrast, among moderate-stenosis lesions, inclusion of CFD-derived features was associated with an improvement beyond using basic CTCA features alone, with earlier divergence of risk curves in model fit compared with basic CTCA features and plaque components and PCAT volumes derived from MEDIS (p=0.007). These findings were supported by analyses of observed cumulative risk with numbers at risk along follow-up, which demonstrated consistent cohort-specific patterns across nested models and are presented in the **Supplementary Figure S2**.

**Figure 4.**
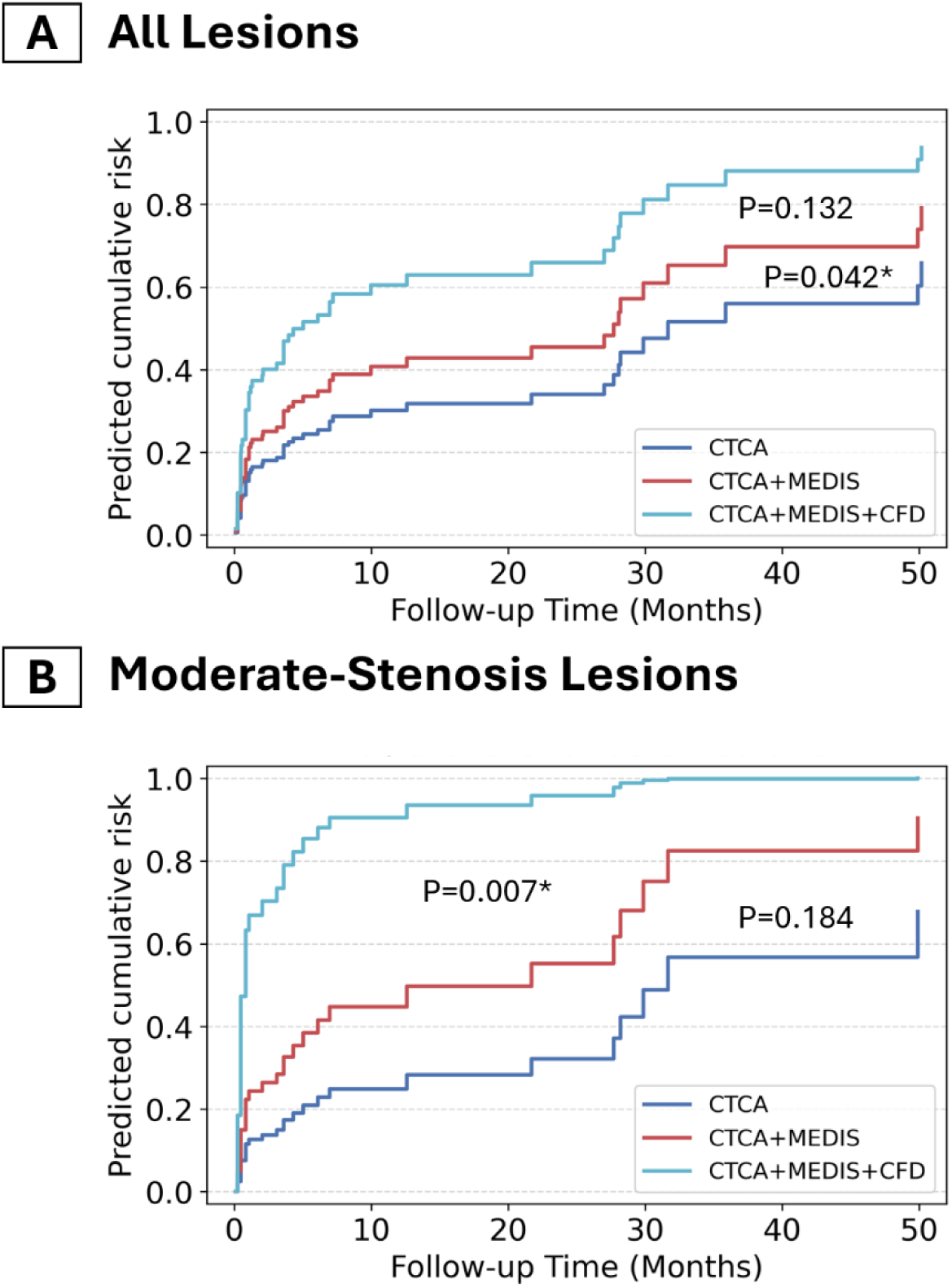
Predicted cumulative risk across nested CT-based models in the overall cohort and the moderate-stenosis subset. Cox model–derived predicted cumulative risk curves are shown for three nested diagnostic strategies: CTCA (anatomical assessment alone), CTCA+MEDIS (addition of quantitative plaque and PCAT metrics), and CTCA+MEDIS+CFD (further incorporation of haemodynamic and geometric features). Left panel depicts all lesions; right panel shows lesions with moderate stenosis. Curves illustrate the stepwise separation of predicted risk with sequential model enrichment. P values annotate pairwise comparisons between nested models (likelihood ratio tests); * denotes statistical significance.

## 4. Discussion

In this within-patient, lesion-level case-control analysis derived from the longitudinal GeoCAD cohort, we found that the imaging determinants of future culprit lesions vary with lesion stenosis severity. Whilst stenosis severity remains the dominant discriminator across the full spectrum of lesions, lesions causing only moderate stenosis exhibited a distinct phenotype where lesion-adjacent PCAT, and CTCA-derived anatomy-flow descriptors (resting haemodynamics and centreline geometry) provided incremental prognostic information beyond luminal narrowing and quantitative plaque composition.

Framed within a CTCA workflow, these findings suggest that advanced post-processing may be most informative when applied selectively to moderate, non–flow-limiting lesions, where stenosis alone offers limited discrimination. This supports integrated CT phenotyping as a risk-enrichment strategy rather than a replacement for established physiology-guided approaches. From an implementation perspective, quantitative plaque and PCAT metrics are increasingly available, whereas CFD-derived haemodynamic and geometric descriptors require additional resources and standardisation. Accordingly, advanced modelling may be selectively applied to moderate lesions following routine CTCA, with future work needed to validate incremental value, reproducibility, and clinical feasibility.

### 4.1 Incremental prognostic value beyond stenosis severity

Coronary stenosis severity has long been central to CT-based assessment of coronary artery disease. When considered across a broad spectrum of lesion severities, %AS serves as an effective surrogate for cumulative plaque burden and overall disease severity, and therefore often used in prediction models that include both mild and severe disease ^20^. Importantly, this observation should be distinguished from clinical studies focusing on intermediate-to-severe lesions, in which invasive physiological indices such as FFR consistently outperform anatomical stenosis in predicting ischaemia and guiding revascularisation ^3^.

In the present study, the contribution of %AS to model discrimination diminished markedly within the moderate-stenosis subgroup (%DS 25–49), where exclusion of %AS resulted in minimal change in Harrell’s C-index. This finding may appear surprising, however, aligns with pathological and longitudinal imaging studies demonstrating that many culprit plaques responsible for acute coronary syndromes arise from angiographically mild-to-moderate lesions, rather than from haemodynamically significant stenoses ^1^. In such lesions, anatomical narrowing alone inadequately captures the biological processes that govern plaque destabilisation, including local inflammation and disturbed haemodynamics.

Historically, both risk stratification and physiological testing have concentrated on angiographically intermediate lesions (commonly 50–70%) to identify flow-limiting disease suitable for revascularisation, leaving earlier moderate lesions relatively underexplored despite their numerical predominance and potential for progression ^22,23^. Our findings extend this concept by providing quantitative lesion-level evidence that, within this earlier disease window, non-anatomical features—specifically pericoronary inflammation, lesion-specific haemodynamic patterns, and vessel geometry—contribute more substantially to culprit discrimination than stenosis severity alone.

### 4.2 Lesion-level PCAT volume as an independent marker of future culprit lesions

Among evaluated features in the moderate-stenosis subgroup, lesion-adjacent PCAT volume contributed the largest independent increment to model discrimination. PCAT is increasingly recognised as an active paracrine tissue that both reflects and modulates vascular inflammation, which may promote progression of atherosclerosis and plaque instability ^8^.

Several factors likely explain why PCAT volume, rather than attenuation, emerged as the dominant signal. First, attenuation studies often rely on proximal-RCA sampling, which may not capture lesion-local inflammatory ^24^. Second, attenuation is sensitive to acquisition parameters introducing variability in lesion-level analyses. Third, PCAT volume provides a spatially localised and stable descriptor of perivascular tissue surrounding the lesion, whereas attenuation-based measures are thought to be more sensitive to dynamic inflammatory states ^25^. Together, these findings support lesion-level PCAT volume as an independent discriminator in moderate stenosis and highlight the need for prospective validation of integrated imaging strategies.

### 4.3 Lesion-specific haemodynamic pattern variations quantified by PCA

Beyond pericoronary inflammation, lesion-specific haemodynamic disturbances and local vessel geometry emerged as key discriminators of future culprit lesions, particularly in the moderate-stenosis subgroup. Our findings that culprit lesions were characterised by greater exposure to high ESS and increased shear stress pattern differences are consistent with a substantial body of experimental and computational literature linking focal high shear, shear gradients, and dynamic shear instability to plaque cap thinning and rupture-prone phenotypes ^5,26^. In this context, the observed association between higher TSVI and future culprit events aligns with recent clinical CFD studies showing that indices capturing cyclic shear expansion–contraction and topological shear variation outperform static or time-averaged shear metrics in identifying myocardial infarction–related lesions^20,27^.

Endothelial cells respond not only to absolute shear but also to spatial and temporal variations, which trigger inflammatory and remodelling pathways ^28^. To characterise these perturbations further at the lesion level, we summarised haemodynamic metrics using PCA across multiple spatial locations along each lesion. To capture lesion-level patterns, haemodynamic metrics were summarised using PCA across spatial locations. This approach reduces collinearity and reflects spatially evolving haemodynamic environments rather than isolated point values.

### 4.4 Complementary roles of flow pattern and vessel geometry in culprit lesion risk

Beyond surface-based shear descriptors, volumetric flow complexity (*h*₂) and centreline geometry (torsion) contributed independently to culprit discrimination. Centreline torsion measures the out-of-plane twisting of the vessel path and, together with curvature, modulates the generation and stability of secondary and helical flow structures ^21^. *h*₂ reflects the degree of three-dimensional rotational flow and captures aspects of flow organisation that influence near-wall transport, supporting an overall atheroprotective role under physiological conditions ^29^.

Although vessel torsion and helical flow are typically positively related, this relationship may be altered by atherosclerotic disease. In this study, culprit lesions showed higher h₂ despite lower torsion, indicating decoupling between vessel geometry and flow topology. This pattern suggests that intensified or disorganised helical flow may arise independently of centreline torsion, potentially driven by local stenosis, plaque morphology, or altered boundary conditions that redistribute flow energy within the lumen ^21,30^. Accordingly, *h*₂ and torsion should be interpreted as complementary descriptors whose combined assessment provides insight into lesion-specific flow–geometry interactions relevant to plaque destabilisation.

### 4.5 Study Limitations

Several limitations should be acknowledged. First, the study lacked an external control group, and culprit and non-culprit lesions were compared within patients. Although this within-patient design reduces confounding variables from fixed patient-level characteristics, such as age, sex, systemic risk factors, and baseline medication exposure, it does not fully account for time-varying determinants of treatment decisions. Second, the primary endpoint was clinically indicated revascularisation, which is influenced by symptoms, downstream testing, and operator judgement; therefore, indication bias cannot be excluded, and these findings should not be interpreted as direct evidence for acute coronary syndrome. Third, the modest sample size and number of events may predispose to model optimism despite feature reduction and bootstrap validation, and external validation is required before clinical application. Fourth, coronary haemodynamics were simulated under resting conditions, without modelling hyperaemia. Hyperaemic simulations are most commonly used for FFR computation to identify flow-limiting stenoses that may benefit from revascularisation. In the present cohort, the majority of lesions were below 50% diameter stenosis and therefore unlikely to demonstrate marked hyperaemia-dependent pressure drops. Accordingly, resting haemodynamic simulations were considered appropriate for characterising lesion-specific flow patterns relevant to plaque biology rather than ischaemia-driven decision-making. Nevertheless, the absence of hyperaemic modelling limits direct comparison with FFR-based studies and may underestimate haemodynamic differences in more advanced lesions. Finally, as with all retrospective imaging studies, causal inference cannot be established, and the findings should be regarded as hypothesis-generating.

## 5. Conclusions

In this lesion-level CTCA study, the determinants of future culprit lesions varied with stenosis severity. While stenosis severity remained influential overall, lesion-adjacent PCAT, volumetric flow topology, and vessel geometry provided incremental prognostic information in moderate-stenosis lesions. Integration of these descriptors improved lesion-level risk discrimination beyond anatomical assessment and quantitative plaque composition, supporting further validation of selective, advanced CT analysis as a risk-enrichment strategy for biologically high-risk, nonobstructive coronary lesions.

## Clinical Perspectives

### Clinical Competencies

In patients undergoing CTCA, many future culprit lesions arise from lesions with only moderate stenosis. In this study, lesion-adjacent PCAT and CT-derived anatomy–flow descriptors (resting haemodynamics and centreline geometry) improved lesion-level discrimination within moderate stenoses beyond anatomical severity and quantitative plaque composition. These findings support competency in quantitative CTCA interpretation and imaging-based risk stratification, and motivate selective use of advanced CTCA analysis to identify moderate lesions that may warrant intensified preventive management.

### Translational Outlook

Next steps include external validation in larger, multicentre cohorts; assessment of reproducibility across scanners and protocols; and comparison with established CT-based approaches (e.g., high-risk plaque features and CT-derived physiology) to define incremental value. Translation will require streamlined, automated workflows with standardisation, quality control, and clinically interpretable thresholds. Prospective studies should evaluate whether integrated CT phenotyping improves risk-enrichment for preventive therapies and reduces downstream events, and how such approaches may be incorporated into clinical decision pathways.

## Funding

S.B was awarded a National Health and Medical Research Council (NHMRC), Australia, Ideas Grant (2012474) and a New South Wales (NSW) Health Cardiovascular Research Capacity Program funding. R.M.G. is supported by a NHMRC L3 Investigator Grant (APP2010203) and a NSW Health Cardiovascular Senior Scientist Grant. L.M-C. acknowledges funding support from a NHMRC Postgraduate Scholarship (GNT2013809) with co-funding from a National Heart Foundation PhD Scholarship (106228), a Royal Australasian College of Physicians Research Establishment Fellowship (2026REF056) and a University of New South Wales Faculty of Medicine and Health Collaborative Seed Grant. The authors acknowledge the NSW Ministry of Health and the Centre for Health Record Linkage for providing data and record linkage services, and Spectrum Medical Imaging for the provision of medical imaging data.

## Supporting information

combined supplemental methods and results

## Data Availability

The data underlying this article cannot be shared publicly due to ethical and patient confidentiality constraints. De-identified data may be available from the corresponding author upon reasonable request and subject to approval by the institutional review board.

## ABBREVIATIONS AND ACRONYMS

ACS: acute coronary syndrome
AS: area stenosis
AUC: area under the curve
CAD: coronary artery disease
CFD: computational fluid dynamics
CI: confidence interval
CTCA: computed tomography coronary angiogram
DS: diameter stenosis
ESS: endothelial shear stress
ESSG: endothelial shear stress gradient
FFR: fractional flow reserve
HR: hazard ratio
LowESS: low endothelial shear stress
HighESS: high endothelial shear stress
MACE: major adverse cardiac event
PC1: first principal component
PCA: principal component analysis
PCAT: pericoronary adipose tissue
ROC: receiver operating characteristic curve
TA-ESS: time-averaged endothelial shear stress
TSVI: topological shear variation index

## Notes

### Competing Interest Statement

The authors have declared no competing interest.

