## Supplementary material for "CTCA-Based Pericoronary Fat and Anatomy–Flow Signatures Identify Future Culprit Lesions in Moderate Stenoses": combined supplemental methods and results

### Supplementary Methods

#### S1. Plaque and Centreline Anatomy Characterisation

Quantitative anatomical descriptors derived from CTCA were defined according to standard coronary imaging conventions and computed using MEDIS QAngio CT (version [insert]) and centreline-based geometric analysis tools (VMTK v1.4). Lesion length was measured as the centreline distance between manually annotated proximal and distal lesion boundaries. Minimal lumen area (MLA) was obtained at the narrowest luminal cross-section. Plaque burden was defined as the proportion of vessel area occupied by plaque at the lesion cross-section (i.e., plaque area divided by vessel area). The remodelling index was defined as lesion vessel area divided by the mean vessel area at proximal and distal reference segments; positive remodelling was classified using a prespecified threshold (e.g., remodelling index >1.10; please confirm threshold used). Eccentricity represented the ratio of maximum to minimum wall thickness within the lesion cross-section. Percentage diameter stenosis (%DS) and area stenosis (%AS) were derived by comparing luminal dimensions at the lesion throat with those of adjacent reference segments. Local geometric descriptors (curvature and torsion) were derived from VMTK-generated centrelines, which provided high-resolution sampling along the vessel path, using standard differential-geometry definitions as shown below.[1]

$$\kappa_a = \frac{1}{\mathrm{L}}\int_{s1}^{s2} \frac{|c'(s) \times c''(s)|}{|\, c'(s)|^{3}} ds, \tag{1}$$

$$\tau_a = \frac{1}{L}\int_{s1}^{s2} \frac{|c'(s) \times c''(s)| \cdot c'''(s)}{|\, c'(s) \times c''(s)|^{2}} ds \tag{2}$$

where c(s) denotes the centreline parameterised along the coordinate s of curve c, and L represents the length of a centreline curve considered.

#### S2. Transient Coronary Blood Flow Simulations

We performed transient coronary blood flow simulations using ANSYS-CFX (v2023r1). Coronary lumen geometries were discretised with unstructured meshes; a mesh-dependence study was used to select the final mesh resolution (maximum element size near the wall: 0.15 mm). Vessel walls were modelled as rigid with a no-slip boundary condition. Inlet flow waveforms at the RCA and LM were derived from literature [2] and applied as time-varying boundary conditions after scaling with the patient-specific vascular inlet diameters with an exponent of 2.55 [3,4]. A scaling law was used to split flow at each bifurcation with an exponent of k = 2.33 [4]. Blood was assumed to be a non-Newtonian fluid using the Carreau–Yasuda model:

$$\mu = \mu_{\mathrm{i}} + \frac{\mu_0 - \mu_{\mathrm{i}}}{[1 + (\lambda|\dot{\gamma}|)^{b}]^{a}} \tag{3}$$

where $\mu$ is the viscosity, $\mu_{\mathrm{I}}$ = 0.0035 Pa·s is the high shear viscosity, $\mu_0$ = 0.16 Pa·s is the low shear viscosity, $\lambda$ = 8.2s is the time constant, and $a$ = 0.64, b = 1.23, following Razavi et al. [5]

Waveforms of the blood flow at the RCA and LM inlets were derived from literature as shown below [2].

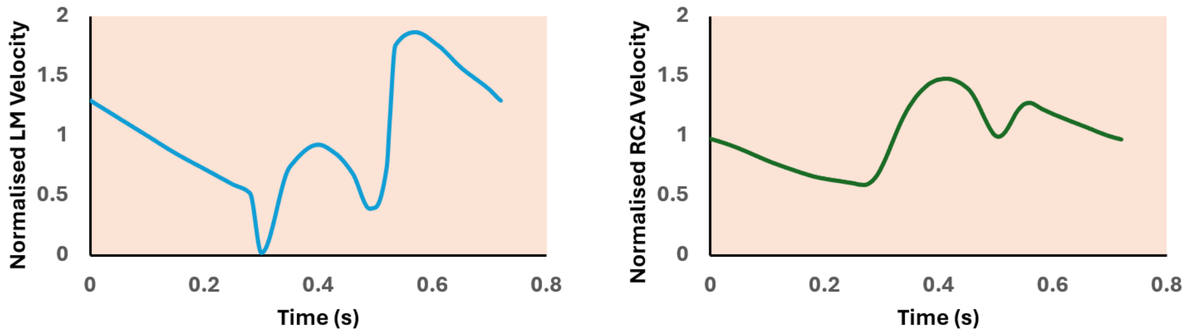


Figure S1. Blood flow velocity conditions specified at the left main (LM, left) and right coronary artery (RCA, right) inlets.

A time step of 0.001 seconds was specified for the implicit second order temporal discretisation scheme within the ANSYS-CFX solver. The criterion for convergence was set as $10^{-4}$ for the continuity and normalised velocity and pressure. We quantified time-average endothelial shear stress (TA-ESS), TA-ESS gradient (TA-ESSG), and helical flow intensity ($h_2$) due to their association with endothelial cell dysfunction [1,6], calculated as:

$$\mathrm{TAESS} = \frac{1}{T}\int_0^T |\tau_\omega| dt \tag{4}$$

$$\mathrm{TAESSG} = \frac{1}{T}\int_0^T |\nabla_s \tau_w| dt \tag{5}$$

$$h_2 = \frac{1}{T}\int_0^T \iiint_V |v(x,t)\cdot\omega(x,t)| dV\, dt \tag{6}$$

where $\tau_w$ is the flow-induced shear stress vector at the luminal wall, T denotes the cardiac cycle period, and and are the velocity field and vorticity at location $x$ and time $t$. In addition, the topological shear variation index (TSVI) that characterises the endothelial contraction and expansion was calculated, following

$$\mathrm{TSVI} = \left[\frac{1}{\mathrm{T}}\int_0^{\mathrm{T}} (\mathrm{DIV_{ESS}} - \overline{\mathrm{DIV_{ESS}}})^2 dt\right]^{\frac{1}{2}} \tag{7}$$

where $\mathrm{DIV_{ESS}}$ is the divergence of the ESS unit vector field, following the definition by Mazzi et al. [7]

Time-averaged endothelial shear stress (TA-ESS) represents the mean tangential force per unit area acting on the endothelium across the cardiac cycle. Low ESS burden was defined as the percentage of luminal surface exposed to ESS <0.5 Pa, a threshold associated with atheroprone biology [8], while High ESS burden represented the percentage of surface exposed to ESS >4.71 Pa, which has been linked to plaque cap thinning and rupture. TA-ESS gradient (TA-ESSG) describes spatial heterogeneity in shear stress and was computed as the spatial derivative of ESS along the vessel surface. TSVI quantifies variations in the shear vector orientation and magnitude throughout the cardiac cycle, capturing dynamic endothelial deformation patterns. Helical flow intensity ($h_2$) reflects the degree of rotational (swirling) flow within the vessel and characterises three-dimensional flow complexity that may not be captured by surface-based metrics alone.

Simulations were run for four cardiac cycles with a time step sufficiently small to resolve pulsatile flow features. Only the final cycle was used for analysis to avoid the influence of initial transients. From the resulting velocity and wall shear stress fields, haemodynamic metrics were sampled at anatomically defined segments across each lesion (proximal, throat, distal) over a 3-mm segment length and over three longitudinal spans, allowing a detailed appraisal of local and evolving flow disturbances. To incorporate the full spatial behaviour of each metric, principal component analysis (PCA) was applied to all location-specific measurements for each variable. The first principal component, consistently capturing the majority of spatial variance along the lesion, was retained as the surrogate descriptor of the overall haemodynamic pattern for subsequent statistical modelling.

### S3. Statistical Analysis

#### S3.1 Data Preprocessing

All lesion-level variables underwent systematic preprocessing prior to formal modelling. Continuous variables were examined for distributional characteristics using histograms, Q–Q plots, and the Shapiro–Wilk test. Variables exhibiting marked skewness or outliers were transformed (logarithmic or rank-based) when appropriate, and all continuous predictors were subsequently standardised to zero mean and unit variance to facilitate comparability across biological domains.

Because haemodynamic and anatomical descriptors were sampled at multiple spatial locations along each lesion, these variables were intrinsically correlated and captured complementary aspects of local versus longitudinal behaviour. To reduce dimensionality while retaining the dominant spatial signature of each metric, principal component analysis (PCA) was performed on the set of location-specific measurements for every haemodynamic and centreline-derived geometric variable. PCA was applied to the standardised data matrix for each metric, and the first principal component (PC1) was retained as the surrogate descriptor of its spatial pattern. PC1 consistently explained the majority of variance and effectively summarised how each metric evolved from proximal to distal segments. This approach mitigated collinearity, reduced the risk of model overfitting, and ensured that lesion-level analyses reflected pattern variation rather than isolated point values, a key consideration for understanding how haemodynamics and geometry interact across the lesion length.

#### S3.2 Feature Selection Through Univariable Screening

Each lesion-level variable was initially screened using univariable Cox proportional hazards models, with lesion follow-up time from the index CT to MACE used as the time scale. Hazard ratios and p-values were estimated for all anatomical, plaque composition, PCAT, geometric, and haemodynamic variables. Proportional hazards assumptions were assessed through visual inspection of Schoenfeld residuals; when violations were identified, variables were either transformed, modelled using an alternative specification (e.g., categorisation), or excluded from multivariable modelling (please confirm which approach was applied).

To construct interpretable and statistically stable multivariable models, variables were organised into six prespecified families representing distinct biological domains: anatomical stenosis severity, plaque composition, PCAT, vessel surface shear metrics, volumetric flow topology, and centreline-based geometry. Within each family, the most prognostically informative variable—defined by the highest HR magnitude or strongest mechanistic justification when HRs were comparable—was selected as the representative feature. This family-based approach prevented the introduction of redundant, collinear predictors; preserved the biological structure of the data; and adhered to event-per-variable constraints inherent to lesion-level time-to-event modelling.

#### S3.3 Incremental Prognostic Value Assessment

Incremental prognostic value was evaluated using a structured modelling strategy designed to quantify the independent contribution of each biological domain beyond anatomical severity. A base Cox model comprising only anatomical stenosis measures was constructed first. Representative features from each of the remaining families were then added sequentially. For each incremental step, model performance was reassessed using changes in Harrell's C-index and likelihood ratio (LR) $\chi^2$ statistics. For drop-one analyses, ΔC-index was computed as the C-index of the full multivariable model minus the C-index of the corresponding model with the feature removed.

This framework allowed formal testing of whether domains such as PCAT, cycle-averaged haemodynamics under resting flow, or volumetric flow complexity contributed predictive information beyond anatomic narrowing. Because PC1-derived haemodynamic descriptors summarised spatial variation across the entire lesion, improvements in model fit were interpreted as evidence that the lesion's longitudinal mechanical environment—rather than isolated shear values—carried prognostic importance. Sensitivity analyses demonstrated that PC1-based metrics provided more robust predictions than using proximal/throat/distal values individually.

A prespecified exploratory analysis was performed for lesions with moderate stenosis (25–49% diameter stenosis) to determine whether incremental prognostic contributions were preserved in lesions that are radiologically non-obstructive but clinically important. Variable selection and modelling procedures remained identical to those used in the overall cohort.

#### S3.4 Clinical Workflow Modelling

To complement the mechanistic framework, a second modelling strategy was constructed to reflect a realistic clinical workflow. Three hierarchical Cox models were developed according to the type and complexity of data available at each stage of the CT diagnostic pathway. The first model incorporated only anatomical stenosis and geometric severity parameters readily available from routine CTCA. The second model added plaque composition and PCAT metrics derived from semi-automated MEDIS analysis, representing contemporary quantitative CT plaque assessment. The third and most comprehensive model included both centreline-derived geometric descriptors and CFD-derived haemodynamic metrics, representing an advanced computational analysis layer.

For each model, predicted risk scores were generated at the lesion level and used to construct cumulative risk curves, illustrating visually how risk stratification improved with the addition of quantitative plaque and haemodynamic information. Time-dependent ROC curves and C-indices at the median follow-up were also computed to compare discrimination across the three workflow models. This hierarchical framing enabled assessment of not only the mechanistic value of each domain but also the incremental **clinical utility** of incorporating more advanced computational analyses into CTCA workflows.

### Supplementary Results

**Table S1**. Blood flow and vessel anatomic characteristics across the lesion length.

| Metrics | Culprit Lesions (n = 55) | Non-culprit Lesions (n = 157) | Adjusted P-values |
|---|---|---|---|
| **TA-ESS, Pa** | | | |
| Proximal | 1.31 (0.9–3.07) | 1.46 (0.9–2.66) | 0.986 |
| Throat | 8.83 (3.75–15.59) | 3.29 (1.54–7.41) | <0.001* |
| Distal | 1.59 (0.7–4.25) | 1.46 (0.84–2.76) | 0.687 |
| PC1 | -0.4 (-0.65–0.47) | -0.59 (-0.72–-0.22) | 0.022* |
| **TA-ESSG, Pa mm$^{-1}$** | | | |
| Proximal | 0.61 (0.38–1.73) | 0.67 (0.36–1.52) | 0.983 |
| Throat | 6.91 (2.37–15.83) | 1.95 (0.69–5.34) | <0.001* |
| Distal | 1.49 (0.4–3.99) | 0.9 (0.42–2.45) | 0.175 |
| PC1 | -0.42 (-0.72–0.41) | -0.68 (-0.82–-0.28) | 0.010* |
| **LowESS, %** | | | |
| Proximal | 0.64 (0.0–21.92) | 2.27 (0.0–13.67) | 0.986 |
| Throat | 0.0 (0.0–0.0) | 0.0 (0.0–3.27) | 0.034* |
| Distal | 5.07 (0.0–42.01) | 7.27 (0.0–28.71) | 0.793 |
| PC1 | -0.73 (-1.2–0.35) | -0.82 (-1.24–0.19) | 0.783 |
| **HighESS, %** | | | |
| Proximal | 0.0 (0.0–7.66) | 0.0 (0.0–10.17) | 0.986 |
| Throat | 71.18 (25.0–91.55) | 16.59 (0.0–66.32) | <0.001* |
| Distal | 5.54 (0.0–32.03) | 0.0 (0.0–15.36) | 0.079 |
| PC1 | 0.08 (-1.24–2.22) | -1.27 (-1.91–0.87) | 0.008* |
| **TSVI, $10^{-3}$ mm$^{-1}$** | | | |
| Proximal | 135.61 (89.65–232.32) | 129.22 (59.99–192.64) | 0.313 |
| Throat | 213.91 (137.42–354.72) | 146.41 (53.7–278.11) | 0.014* |
| Distal | 431.22 (269.57–556.63) | 218.47 (112.74–451.82) | 0.002* |
| PC1 | 1.16 (-0.51–2.02) | -0.88 (-1.8–1.11) | 0.002* |
| **Curvature, $10^{3}$ mm$^{-1}$** | | | |
| Proximal | 0.19 (0.15–0.24) | 0.19 (0.15–0.24) | 0.831 |
| Throat | 0.21 (0.17–0.27) | 0.19 (0.14–0.24) | 0.141 |
| Distal | 0.18 (0.15–0.22) | 0.17 (0.13–0.21) | 0.200 |
| PC1 | 0.1 (-0.87–1.25) | -0.48 (-1.69–0.91) | 0.107 |
| **Torsion, mm$^{-1}$** | | | |
| Proximal | 18.64 (11.49–21.95) | 21.51 (14.73–34.35) | 0.033* |
| Throat | 14.29 (11.81–19.81) | 18.71 (12.57–31.01) | 0.009* |
| Distal | 15.74 (11.82–21.09) | 19.29 (14.03–28.18) | 0.013* |
| PC1 | -0.85 (-1.42–-0.29) | -0.3 (-1.18–1.16) | 0.009* |
| **$h_2$** | | | |
| Proximal | 12.28 (4.07–55.81) | 6.27 (2.4–15.39) | 0.009* |
| Throat | 18.82 (4.66–73.81) | 7.6 (2.51–21.51) | 0.005* |
| Distal | 24.01 (6.08–184.1) | 7.91 (2.7–29.7) | 0.005* |
| PC1 | -0.27 (-0.36–0.03) | -0.34 (-0.37–-0.27) | 0.004* |

Note: Values are given as median (interquartile ranges). P-values are from Mann–Whitney U tests and were adjusted using the Benjamini–Hochberg false discovery rate procedure. TA-ESS = time-averaged endothelial shear stress; TA-ESSG = time-averaged endothelial shear stress gradient; LowESS = % lumen area exposed to low ESS (<0.5 Pa); HighESS = % lumen area exposed to high ESS (>4.71 Pa); TSVI = topological shear variation index; $h_2$ = helical flow intensity. * indicates statistical significance.

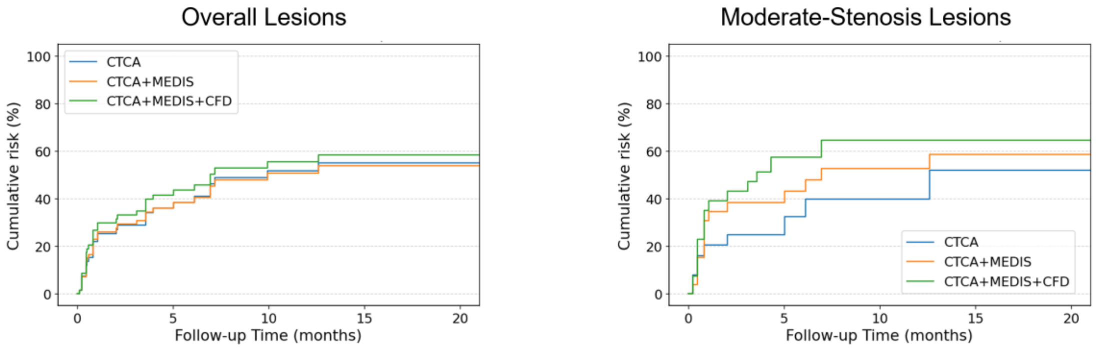


Number at risk (months)

| | 0 | 5 | 10 | 15 | 20 |
|---|---|---|---|---|---|
| CTCA | 70 | 26 | 15 | 9 | 9 |
| CTCA+MEDIS | 70 | 28 | 16 | 10 | 10 |
| CTCA+MEDIS+CFD | 70 | 27 | 15 | 9 | 9 |

Number at risk (months)

| | 0 | 5 | 10 | 15 | 20 |
|---|---|---|---|---|---|
| CTCA | 27 | 10 | 5 | 2 | 2 |
| CTCA+MEDIS | 27 | 13 | 8 | 5 | 5 |
| CTCA+MEDIS+CFD | 27 | 6 | 2 | 2 | 2 |

**Figure S2**. Observed cumulative risk across nested CT-based models in the overall cohort and the moderate-stenosis subset. Kaplan–Meier–derived observed cumulative risk curves are shown for three nested CT-based modelling strategies: CTCA (anatomical assessment alone), CTCA+MEDIS (addition of quantitative plaque and PCAT metrics), and CTCA+MEDIS+CFD (further incorporation of haemodynamic and vascular anatomic features). Left panel displays results for all lesions, while the right panel shows lesions with moderate stenosis. Curves represent the cumulative incidence of culprit-related events over follow-up. Tables below each plot report the number of lesions at risk at prespecified time points. Together, these plots illustrate the progressive separation of observed event risk with stepwise enrichment of CT-derived lesion information.
